# A systematic review of COVID-19 vaccine efficacy and effectiveness against SARS-CoV-2 infection and disease

**DOI:** 10.1101/2021.09.17.21263549

**Authors:** Melissa M Higdon, Brian Wahl, Carli B Jones, Joseph G Rosen, Shaun A Truelove, Anurima Baidya, Anjalika A Nande, Parisa A ShamaeiZadeh, Karoline K Walter, Daniel R Feikin, Minal K Patel, Maria Deloria Knoll, Alison L Hill

## Abstract

Billions of doses of COVID-19 vaccines have been administered globally, dramatically reducing SARS-CoV-2 incidence and severity in some settings. Many studies suggest vaccines provide a high degree of protection against infection and disease, but precise estimates vary and studies differ in design, outcomes measured, dosing regime, location, and circulating virus strains. Here we conduct a systematic review of COVID-19 vaccines through February 2022. We included efficacy data from Phase 3 clinical trials for 15 vaccines undergoing WHO Emergency Use Listing evaluation and real-world effectiveness for 8 vaccines with observational studies meeting inclusion criteria. Vaccine metrics collected include protection against asymptomatic infection, any infection, symptomatic COVID-19, and severe outcomes including hospitalization and death, for partial or complete vaccination, and against variants of concern Alpha, Beta, Gamma, Delta, and Omicron. We additionally review the epidemiological principles behind the design and interpretation of vaccine efficacy and effectiveness studies, including important sources of heterogeneity.

## Introduction

SARS-CoV-2’s rapid global spread and alarming clinical severity have accelerated demand for vaccines that safely and effectively prevent disease or reduce severity. To date, 30 COVID-19 vaccines have received emergency use authorization in at least one country, and nearly 5 billion people have been vaccinated^1,2^.

Evidence from clinical trials and observational studies overwhelmingly supports the safety and efficacy/effectiveness of numerous COVID-19 vaccines, especially against severe disease and death in fully-vaccinated individuals. However, precise estimates of vaccine efficacy and effectiveness have varied across studies due to a variety of factors. For example, efficacy against symptomatic COVID-19 for AstraZeneca’s two-dose viral vector vaccine (AZD1222) ranged from 62—90% in Phase 3 clinical trials^3^, attributed to differences in dosing schedules. Observational studies in a variety of settings also produced a wide range of effectiveness estimates (50—100%) against different clinical outcomes^4–14^. The emergence of SARS-CoV-2 “variants of concern”, including Alpha (B.1.1.7), Beta (B.1.351), Gamma (P.1), Delta (B.1.617.2), and Omicron (B.1.1.529) further confounds interpretation of estimates obtained from comparably-designed vaccine studies. Some variants are associated with higher viral load^15–19^, evasion of neutralizing antibodies *ex vivo*^20–25^, and lower vaccine effectiveness^13,19,26–29^. However, separating diminished protection against a variant from the effects of waning immunity, study methodology, or other contextual factors (e.g., lower use of non-pharmaceutical interventions) is complicated by their co-occurrence and endogeneity.

Previous reviews of COVID-19 vaccines have focused mainly on the developmental pathway and early results from pre-clinical and clinical trials, the immunological basis of vaccine-induced protection, a narrow subset of vaccines or clinical outcomes, or have been rapid in nature and limited to earlier studies^30–47^. There remains an urgent need for comprehensive, up-to-date review of COVID-19 vaccine effects, including real-world evidence. Here, we systematically reviewed COVID-19 vaccine efficacy and effectiveness data against multiple clinical outcomes, for both full and partial immunization courses, by circulating SARS-CoV-2 variants of concern. We focused on studies conducted in primarily adult populations, for the primary vaccine course only (i.e., excluding “booster” doses), and outcomes measured within the first six months after the final vaccine dose.

## Methods

We reviewed studies reporting vaccine *efficacy* (from randomized clinical trials) or *effectiveness* (from observational studies)^48,49^ for vaccines which had received or submitted applications for Emergency Use Listing from WHO as of February 1, 2021^50^ and had at minimum publicly-released data from completed Phase 3 trials (**Table 1**). We searched for clinical trial results published in peer-reviewed scientific journals (via PubMed) and preprint servers (medRxiv, bioRxiv, SSRN), government public health, regulatory agency, and vaccine manufacturers’ websites, and in news articles (Google). Searches were conducted using the vaccine’s brand, trade, or research name. For observational studies, we applied detailed queries to multiple databases (**<u>Supplementary Methods</u>**), and only included results that appeared in at least a detailed report/preprint. Study title and abstract were screened before progressing to full-text review (**<u>Figure S1</u>**). We extracted vaccine efficacy/effectiveness against disease endpoints, specifically asymptomatic infection, any infection, symptomatic disease, severe disease (including “hospitalization”), and death (**<u>Table S1</u>**).

**Table 1:** Status of COVID-19 vaccines within the World Health Organization Emergency Use Listing evaluation process. Vaccine products are listed in descending order based on the number of countries in which the vaccine is approved for emergency use. Vaccines included in the current study - based on availability of efficacy data from Phase 3 clinical trials - are highlighted in grey. Abbreviations: EUL = emergency use listing, EOI = expression of interest, CI = confidence interval. Vaccine details were obtained from McGill University’s COVID-19 Vaccine Tracker: https://covid19.trackvaccines.org/, which aggregates data from multiple sources. Original source for WHO EUL status : https://extranet.who.int/pqweb/key-resources/documents/status-covid-19-vaccines-within-who-eulpq-evaluation-process

| Vaccine Name |  | Vaccine type | WHO Status | Countries |  | Phase 3 Efficacy Trial |  |  | Ref |
| --- | --- | --- | --- | --- | --- | --- | --- | --- | --- |
|  |  |  |  | Developed in | # approved | Data? | CIs? | Peer reviewed? |  |
| **AstraZeneca/Oxford** | AZD1222<br>ChAdOx1-nCoV-19<br>Vaxzevria<br>Covishield | viral vector | EUL Authorized | UK | 162 | Yes | Yes | Yes | 3,68,82,86 |
| **Pfizer/BioNTech** | BNT162b2<br>Comirnaty<br>Tozinameran | mRNA | EUL Authorized | Germany | 134 | Yes | Yes | Yes | 64,71,88 |
| **Janssen/Johnson & Johnson** | Ad26.COV2.S | viral vector | EUL Authorized | USA | 105 | Yes | Yes | Yes | 66 |
| **Sinopharm-Beijing** | BBIBP-CorV<br>Covilo | inactivated virus | EUL Authorized | China | 87 | Yes | Yes | Yes | 89 |
| **Moderna** | mRNA-1273<br>Spikevax | mRNA | EUL Authorized | USA | 85 | Yes | Yes | Yes | 67 |
| **Gamaleya Institute** | Sputnik V<br>Gam-COVID-Vac | viral vector | Submission in Progress | Russia | 74 | Yes | Yes | Yes | 72 |
| **Sinovac** | CoronaVac | inactivated virus | EUL Authorized | China | 52 | Yes | Yes | Yes | 73,74,90 |
| **Novavax** | NVX-CoV2373<br>Nuvaxovid<br>Covovax | protein subunit | EUL Authorized | USA | 31 | Yes | Yes | Yes | 70,83,223 |
| **Bharat Biotech** | BBV152<br>Covaxin | inactivated virus | EUL Authorized | India | 13 | Yes | Yes | Yes | 65 |
| **CanSinoBIO** | Ad5-nCoV<br>Convidecia | viral vector | EOI Accepted | China | 10 | Yes | Yes | Yes | 75 |
| **BioCubaFarma** | Abdala<br>CIGB-66 | protein subunit | Submission in Progress | Cuba | 6 | Yes | Yes | No | 91 |
| **BioCubaFarma** | Soberana 02<br>FINLAY-FR-2 | conjugate | Submission in Progress | Cuba | 4 | Yes | Yes | No | 92 |
| **Vector Institute** | EpiVacCorona | protein subunit | Submission in Progress | Russia | 4 | No | No | No |  |
| **Anhui Zhifei Longcom** | ZF2001<br>ZIFIVAX | protein subunit | EOI Accepted | China | 3 | Yes | No | No |  |
| **Sinopharm-Wuhan** | WIBP-CorV | inactivated virus | EOI Accepted | China | 2 | Yes | Yes | Yes | 89 |
| **IMBCAMS** | Covidful | inactivated virus | Submission in Progress | China | 1 | No | No | No |  |
| **BioCubaFarma** | Soberana 02+<br>FINLAY-FR-1A | protein subunit | Submission in Progress | Cuba | 1 | Yes | Yes | No | 92 |
| **Clover** | SCB-2019 | protein subunit | EOI Accepted | China | 0 | Yes | Yes | Yes | 76 |
| **CureVac** | CVnCoV | mRNA | EOI Accepted/<br>Withdrawn | Germany | 0 | Yes | Yes | Yes | 224 |
| **Sanofi** | CoV2 preS dTM-AS03<br>Vidprevtyn | protein subunit | EOI Accepted | France | 0 | No | No | No |  |

Observational studies were only eligible if the comparison group included concurrent individuals (e.g., modeled or historic controls were excluded), outcomes were laboratory-confirmed, the study design attempted to account for confounding, vaccination status was determined by self-report for ⩽10% of participants, confidence intervals were reported, no significant bias was present as determined by expert opinion, controls were unvaccinated (e.g., excluded if “unvaccinated” included days 0-12 post-vaccination). Only studies comparing persons with and without the clinical outcome under investigation and with and without vaccination were included. Impact studies and studies evaluating progression to severe disease among SARS-CoV-2-positive individuals were excluded. For full immunization (1 or 2 doses, depending on the vaccine), results were included if at least 1 week had passed between the final dose and case detection; for partial immunization, cases must have occurred at least 2 weeks after dose 1 but before dose 2. We excluded efficacy/effectiveness values for which i) follow-up period after final dose was >6 months, ii) doses beyond the primary series were given (i.e., boosters), or iii) multiple vaccines were combined. We classified efficacy/effectiveness against specific SARS-CoV-2 variants if sequencing (or other molecular methods) either i) confirmed the variant in all cases contributing to the estimate or ii) confirmed the variant caused the vast majority of cases in sample of study participants or of the larger population from which they came. Our data are available on *VIEW-hub*, a weekly-updated resource developed by Johns Hopkins’ International Vaccine Access Center (<u>view-hub.org/resources</u>) and in **<u>Table S1</u>**.

## Epidemiological Principles

### Measuring how well a vaccine prevents infection and disease

Vaccine efficacy is evaluated in randomized controlled trials (RCTs) and is defined as the relative reduction in the probability of developing disease in a particular time period in vaccinated individuals compared to unvaccinated individuals. In RCTs, subjects are randomly assigned to receive the vaccine or not (instead usually receiving a placebo or another vaccine). Vaccine efficacy (VE) is calculated using the formula:

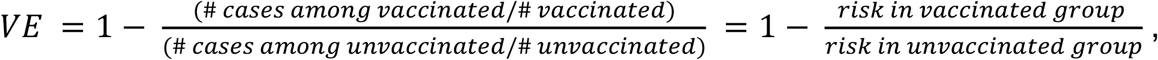

where sometimes the denominator “# vaccinated” (“# unvaccinated”) is replaced with the sum of the total time enrolled in the study among vaccinated (unvaccinated) subjects (i.e., the “person time”)^51,52^. While RCTs are the gold-standard for vaccine studies^53^, they are costly and generally too small to evaluate rare outcomes (e.g., death).

Vaccine efficacy describes the *relative*, as opposed to *absolute*, risk of disease. This is a desired feature of a metric for vaccine strength, since absolute risk may change over time as the background disease incidence changes during an epidemic due to factors like seasonality and behavior change. For example, if the absolute risk of disease in one setting is reduced from 50% to 10% through vaccination, then the vaccine efficacy (80%) is the same as in another setting where the absolute risk was reduced from 5% to 1%. The value of vaccine efficacy also does not tell us whether vaccine failure occurs in a “leaky” or “all-or-nothing” way^52,54,55^.

Once a vaccine is shown to be safe and efficacious in clinical trials and is authorized for general use, further RCTs to evaluate efficacy under new conditions or in special populations are often considered unethical or impractical, especially in the setting of a wide-spread epidemic, because it necessitates withholding vaccines from people who might benefit from them. Instead, observational studies evaluating real-world effectiveness are used to augment efficacy trials, and include designs such as case-control studies (including test-negative designs) and cohort studies (prospective or retrospective)^52,56,57^. Although observational studies must carefully address biases due to differences in those who chose to or were eligible to receive vaccines compared to those who did not, they may provide a more realistic picture of population heterogeneity and include more high-risk groups compared to RCTs. During massive vaccination campaigns like those occurring for COVID-19, observational studies can have much larger sample sizes.

To measure vaccine efficacy/effectiveness, the specific clinical outcome that the vaccine is meant to prevent must be carefully defined. The ideal goal of vaccination is to completely prevent infection (and thus disease and transmission), meaning that vaccine-induced immunity blocks the earliest attempts of the pathogen to replicate within the body. This “sterilizing immunity” is rare, and vaccine efficacy/effectiveness against infection is difficult to measure^58,59^ for short-lived and commonly asymptomatic infections like SARS-CoV-2 since it requires frequent testing of the study cohort. To reduce the public health impact of an infectious disease, prevention of severe disease (including hospitalization and death) is desired, even if infection still occurs^53,59,60^. But because most individuals with COVID-19 recover completely with only mild or moderate symptoms^61–63^, studies of severe outcomes can require hundreds of thousands of participants or months to years of observation time. For COVID-19, the primary endpoint for most clinical trials was symptomatic, laboratory-confirmed COVID-19 disease,^53^ defined as the occurrence of COVID-19-associated symptoms (e.g., cough, shortness of breath, fever) in the presence of detectable SARS-CoV-2. This outcome choice represents a trade-off between public health importance and practicality. Symptomatic disease is on the spectrum leading to severe disease but much more common, and testing can be restricted to those self-reporting symptoms. Although some trials reported efficacy against severe disease as secondary outcomes despite small numbers^64–76^, large observational studies provide more precise estimates.

### Sources of heterogeneity across studies

COVID-19 vaccine studies were conducted by many independent research teams in diverse epidemic settings around the world (**Table 1, Table 2, <u>Table S1</u>, Figure 1**). Consequently, there are several potential sources of heterogeneity between studies that make comparing vaccine efficacy/effectiveness estimates difficult^57,77^:

- <u>Study population</u>: Efficacy/effectiveness can be lower in studies including more participants at high risk of disease (e.g., comorbid individuals) or with reduced immune function (e.g., people living with HIV).
- <u>Outcome and case definition</u>: Vaccine efficacy/effectiveness differs between disease outcomes with varying severity. Even when studies have the same stated outcome, the case definition can vary substantially. For example, most clinical trials used ‘symptomatic COVID-19 disease’ as the primary outcome but included anywhere from 5 (for AstraZeneca/AZD1222^3^) to 16 (Janssen/Ad26.COV2.S^66^) different potential symptoms and varied in requiring one or two to be present. These differences are exacerbated in effectiveness studies that often rely on passive surveillance by health systems. In addition, different definitions in the timing of the disease outcome (e.g., death within 30 days after diagnosis) can lead to heterogeneities.
- <u>Follow-up period</u>: Since developing an adaptive immune response after vaccination takes time^59,78–80^, studies that begin the follow-up period sooner could observe lower efficacy/effectiveness (e.g., 7+ days for Novavax/NVX-CoV2373^70^ vs 28+ days for Janssen/Ad26.COV2.S^66^). In addition, if immune protection wanes, studies with longer follow-up could have lower estimates.
- <u>Predominant variants</u>: Some SARS-CoV-2 variants of concern have been observed to exhibit immune-escape properties^21,81^ (e.g., Beta^28,82–84^, Delta^13,19,65^, Omicron^26,27,29^). Studies conducted when such variants account for a large proportion of infections could have lower efficacy/effectiveness (**Figure 1**).
- <u>Study design and analysis</u>: To make up for lack of randomization, vaccine effectiveness studies can attempt to reduce confounding through study design (e.g., the test-negative design) and during analyses (e.g., controlling for potential confounding variables in regression models). Some studies, especially those that use administrative data, may not collect and therefore cannot control for such possible confounders.

**Table 2.** Summary of vaccine effectiveness studies. Number of included studies by COVID-19 vaccine and by outcome, population, study design, number of doses, and variant of concern. No studies reported for other vaccine candidates met our inclusion criteria (see Methods). Details of all the individual studies are included in **<u>Table S1</u>**.

|  | Vaccine |  |  |  |  |  |  |  | All |
| --- | --- | --- | --- | --- | --- | --- | --- | --- | --- |
|  | BNT162b2<br>(Pfizer-BioNTech) | mRNA-1273<br>(Moderna) | AZD1222<br>(AstraZeneca-Oxford) | Ad26.COV2.S<br>(Janssen/<br>Johnson &<br>Johnson) | Sputnik V<br>(Gamaleya) | CoronaVac<br>(Sinovac) | BBV152<br>(Bharat) | BBIBP-CorV<br>(Sinopharm-Beijing) |  |
| **Total** | 86 | 39 | 30 | 16 | 1 | 6 | 1 | 2 | 107 |
| **Outcome type** |  |  |  |  |  |  |  |  |  |
| Death | 12 | 6 | 5 | 6 | 1 | 4 | 0 | 1 | 21 |
| Severe disease | 38 | 19 | 12 | 10 | 0 | 3 | 0 | 1 | 50 |
| Symptomatic disease | 28 | 12 | 12 | 4 | 0 | 3 | 1 | 0 | 38 |
| Asymptomatic infection | 5 | 3 | 1 | 0 | 0 | 0 | 0 | 0 | 9 |
| Any infection | 58 | 25 | 14 | 7 | 1 | 2 | 0 | 1 | 67 |
| **Population type** |  |  |  |  |  |  |  |  |  |
| General population | 40 | 26 | 15 | 9 | 1 | 2 | 0 | 2 | 45 |
| Health care workers | 18 | 4 | 1 | 1 | 0 | 0 | 1 | 0 | 20 |
| Older adults<br>(≥ 60 years) | 8 | 0 | 8 | 1 | 0 | 2 | 0 | 0 | 11 |
| Other age groups | 6 | 3 | 3 | 2 | 0 | 0 | 0 | 0 | 7 |
| Contacts of cases | 6 | 2 | 3 | 2 | 0 | 0 | 0 | 0 | 6 |
| LTCF/SNF residents | 5 | 1 | 1 | 0 | 0 | 0 | 0 | 0 | 5 |
| Other groups | 6 | 3 | 1 | 1 | 0 | 2 | 0 | 0 | 9 |
| **Study design** |  |  |  |  |  |  |  |  |  |
| Test-negative case control | 30 | 21 | 13 | 7 | 0 | 3 | 1 | 1 | 42 |
| Traditional case-control | 4 | 1 | 0 | 0 | 0 | 0 | 0 | 0 | 3 |
| Prospective cohort | 17 | 4 | 7 | 1 | 0 | 1 | 0 | 0 | 21 |
| Retrospective cohort | 36 | 13 | 9 | 8 | 1 | 2 | 0 | 1 | 41 |
| **# of doses** |  |  |  |  |  |  |  |  |  |
| Complete | 76 | 36 | 19 | 16 | 1 | 6 | 1 | 2 | 96 |
| Partial | 44 | 14 | 25 | N/A | 0 | 3 | 1 | 0 | 53 |
| **Variants of concern** |  |  |  |  |  |  |  |  |  |
| Alpha | 33 | 8 | 17 | 2 | 1 | 0 | 1 | 0 | 35 |
| Beta | 4 | 2 | 1 | 1 | 0 | 0 | 0 | 0 | 6 |
| Gamma | 2 | 2 | 3 | 1 | 0 | 1 | 0 | 0 | 5 |
| Delta | 22 | 13 | 9 | 5 | 0 | 1 | 1 | 0 | 29 |
| Omicron | 4 | 4 | 0 | 0 | 0 | 0 | 0 | 0 | 5 |

**Figure 1.**
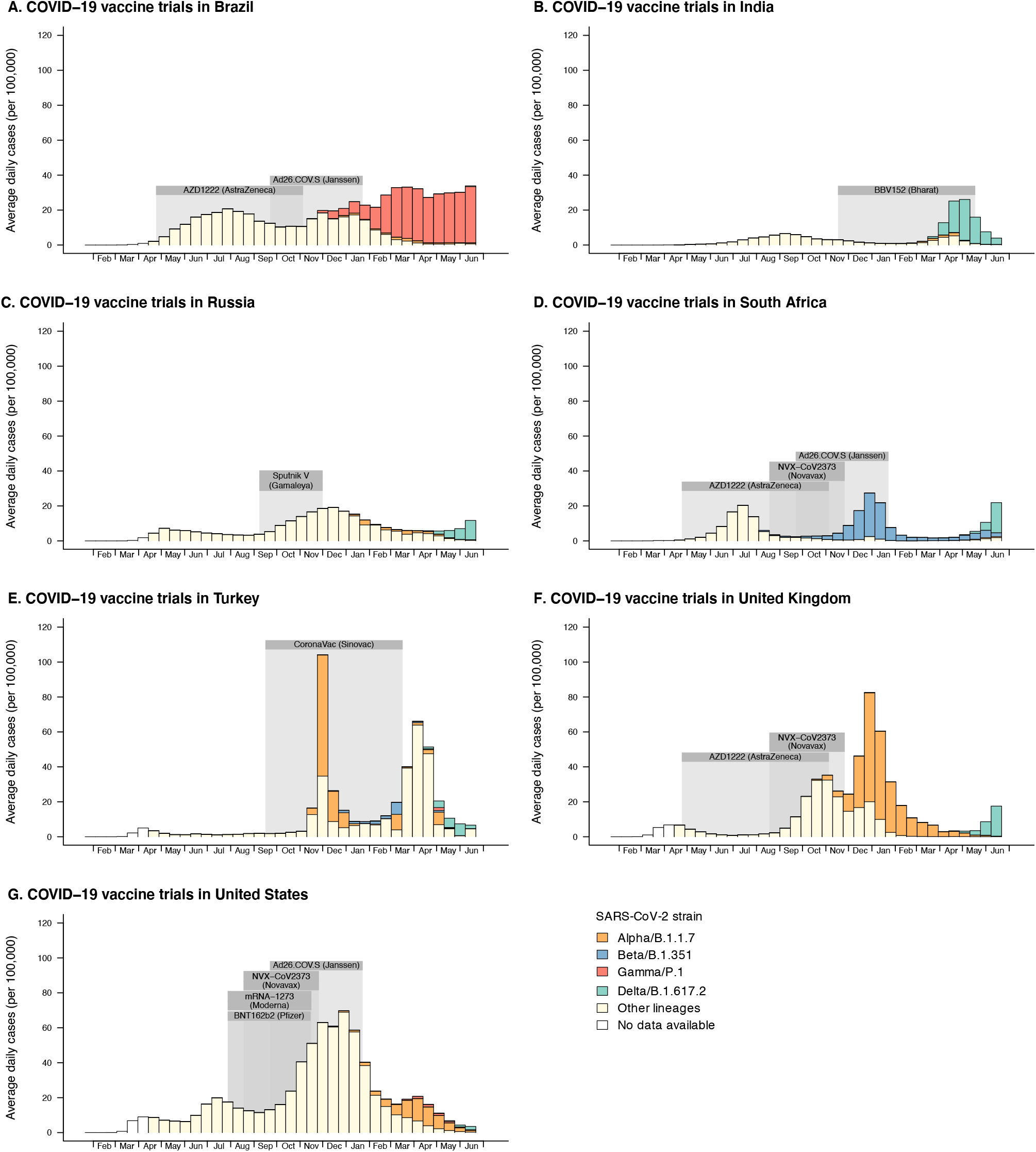
Local context of Phase 3 clinical trials of COVID-19 vaccines. For each country, the time period during which outcomes were observed during each vaccine trial is shaded grey. For each two week period, the average daily incidence of reported cases is shown (height of bars)^225^. The contribution of each major variant of concern to total case counts is estimated from the reported fraction of sequenced SARS-CoV-2 samples belonging to that strain (fill color)^226^. Figure includes only vaccine trials described in published or pre-print reports, trial sites with at least 10,000 individuals from the general adult population, and countries regularly reporting SARS-CoV-2 lineages to the GISAID database.

Other reviews of COVID-19 vaccine efficacy/effectiveness have tended to summarize results from studies with different designs, participant pools, and settings with a single effectiveness value or range, sometimes via formal meta-analysis^36,37,39,41,42,44,47,85^. Here we have taken a different approach, showing individual efficacy/effectiveness measures and their sources, and discussing reasons for the observed heterogeneity, while also highlighting the consensus that emerges among them.

## Results of COVID-19 Vaccine Studies

### Data available from clinical trials of vaccine efficacy

As of Feb 3, 2021, 20 unique vaccine products were in the WHO Emergency Use Listing (EUL) evaluation process, including mRNA, viral vector, inactivated virus, protein subunit, and conjugate vaccine platforms, and were developed by a mix of pharmaceutical companies, non-profit research institutes, and government agencies (**Table 1**). Eight vaccines representing four platforms had received authorization. Efficacy estimates with uncertainty intervals from Phase 3 clinical trials were publicly available for 15 of the 20 vaccine candidates in 22 separate reports^3,64–70,72–76,82,83,86–92^(**Figure 2, <u>Figure S1</u>)**. Vaccine efficacy against symptomatic COVID-19 was available for all 15 vaccines, ranging from 58-95%, with highest efficacy (>90%) from six products representing mRNA, protein subunit and viral vector platforms. Efficacy against any infection was available for five EUL authorized vaccines and against severe disease for six, though confidence intervals for severe disease tended to be wide due to limited sample sizes and trial durations (**Figures 3-7, <u>Table S1</u>**).

**Figure 2.**
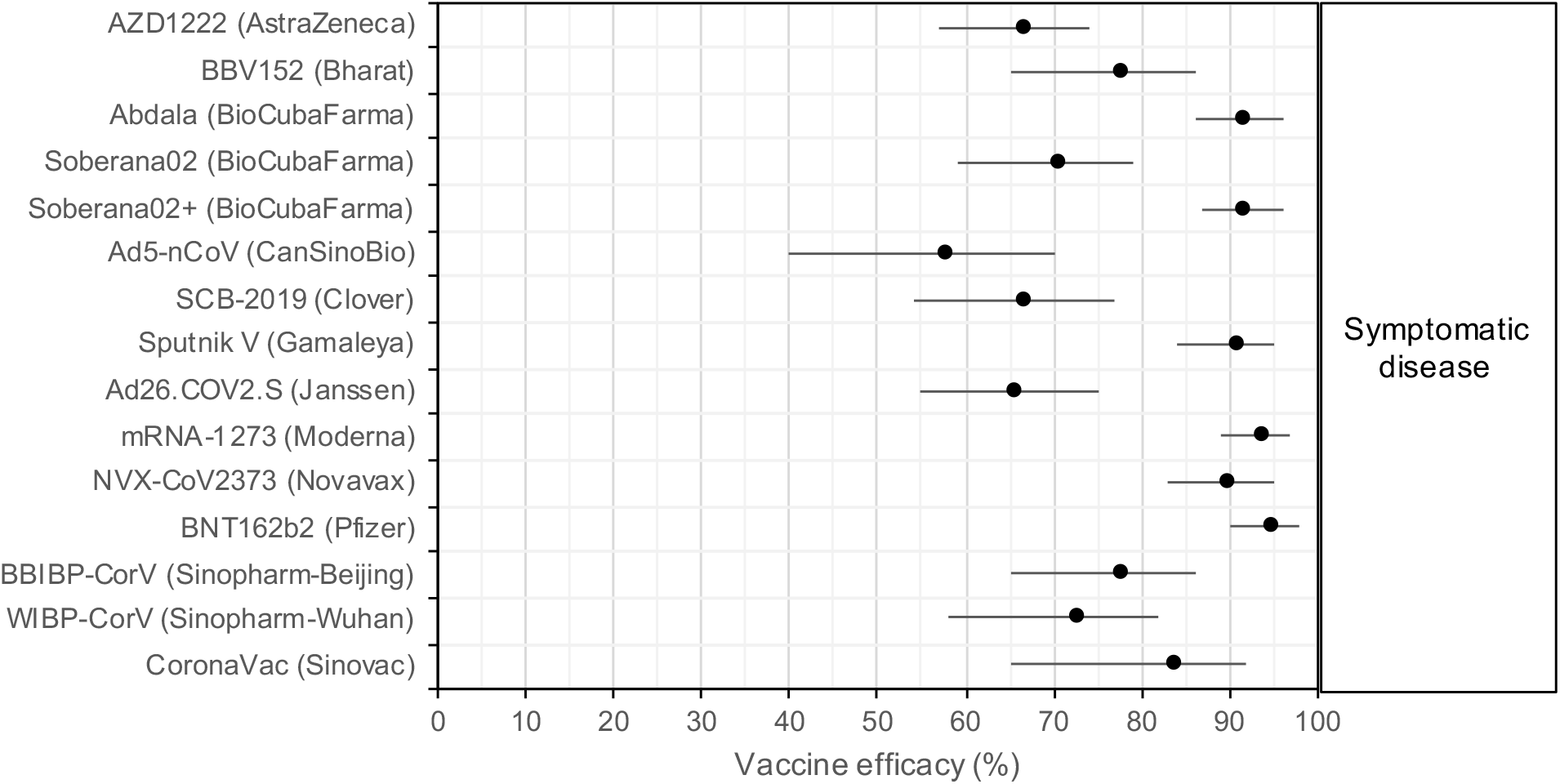
Vaccine efficacy against symptomatic COVID-19, from Phase 3 clinical trials. Each efficacy value is for the complete vaccine course (1 dose for Ad26.COV2.S/Janssen and Ad5-nCoV/CanSinoBio, 3 doses for BioCubaFarma/Abdala and Soberana02+/BioCubaFarma, and 2 doses for all others). Two vaccines which withdrew from the WHO Emergency Use Listing evaluation process but completed Phase 3 trials were not included (CureVac’s mRNA vaccine CVnCoV, reporting 48%

**Figure 3.**
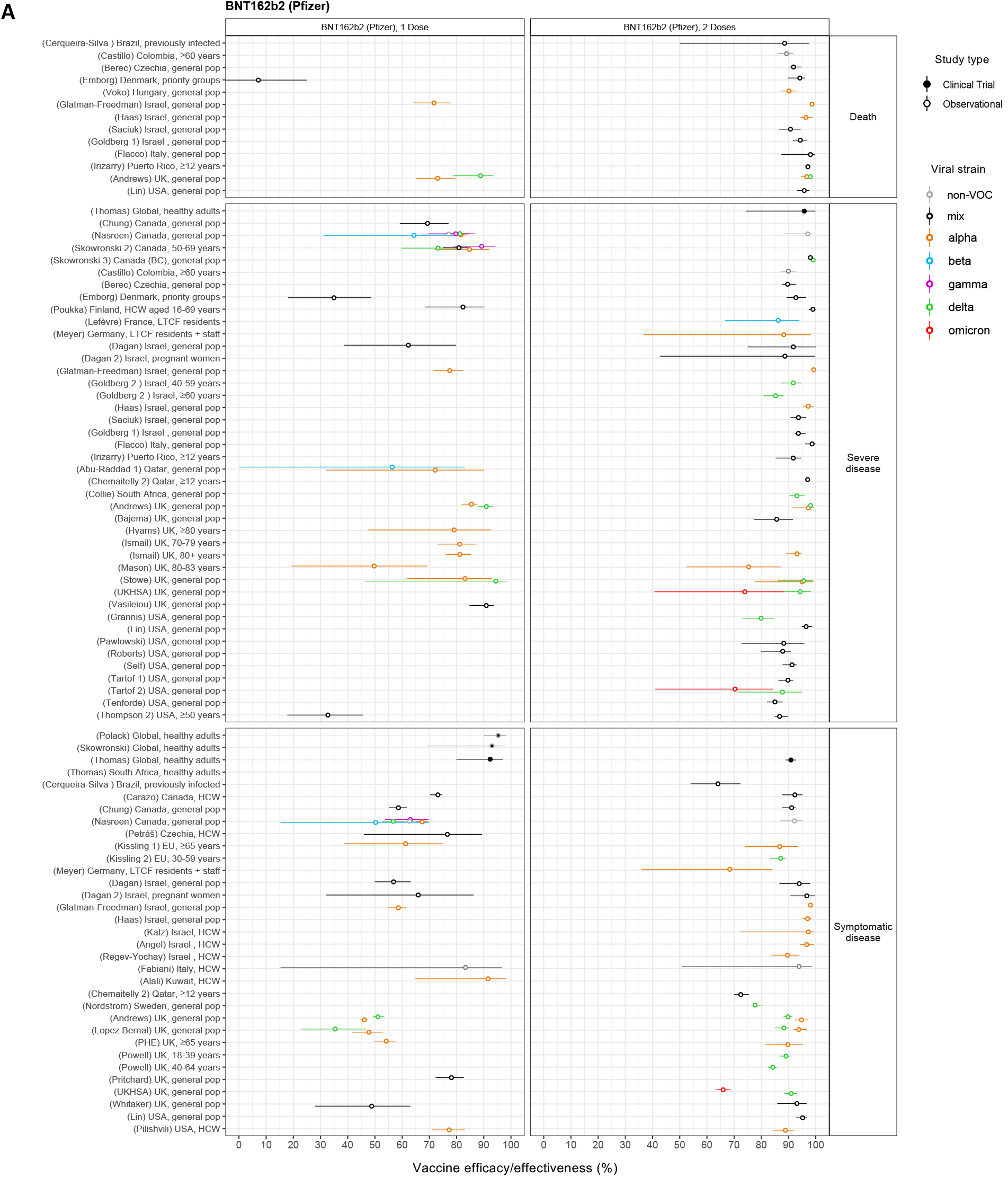
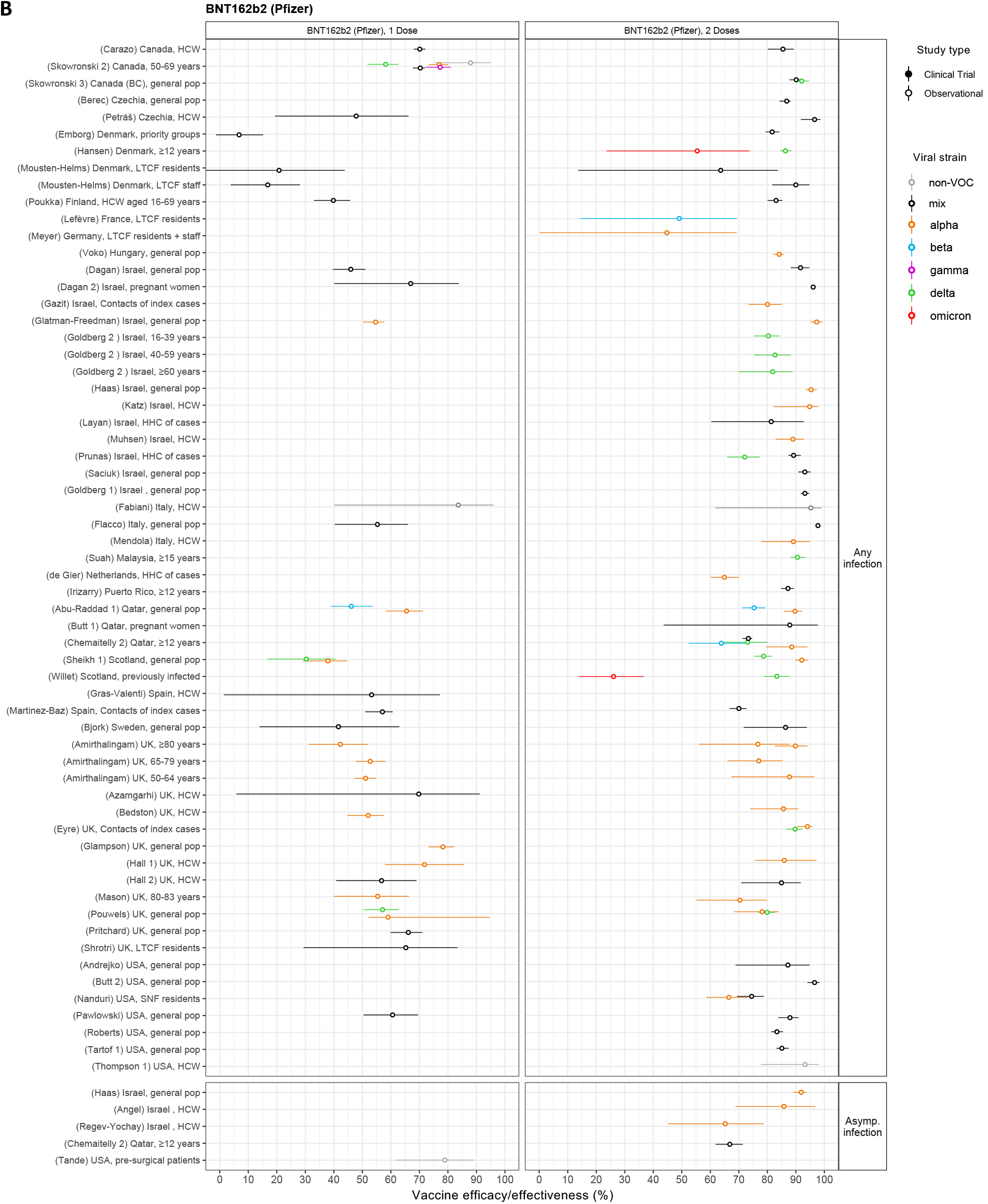
Vaccine efficacy and effectiveness estimates for BNT162b2, a two-dose mRNA vaccine developed by Pfizer/BioNTech. **A)** Efficacy/effectiveness against death, severe disease, or symptomatic disease. **B)** Efficacy/effectiveness against any or asymptomatic infection. Estimates are colored by the viral variant against which the vaccine efficacy or effectiveness value was measured. Solid markers are estimates from randomized clinical trials (efficacy values), and open markers are estimates from observational studies (effectiveness values). The source of each estimate is given by the labels on the left side (“(reference number) Country, population”). Within each disease severity level, estimates are ordered alphabetically by country, and then by population.

**Figure 4.**
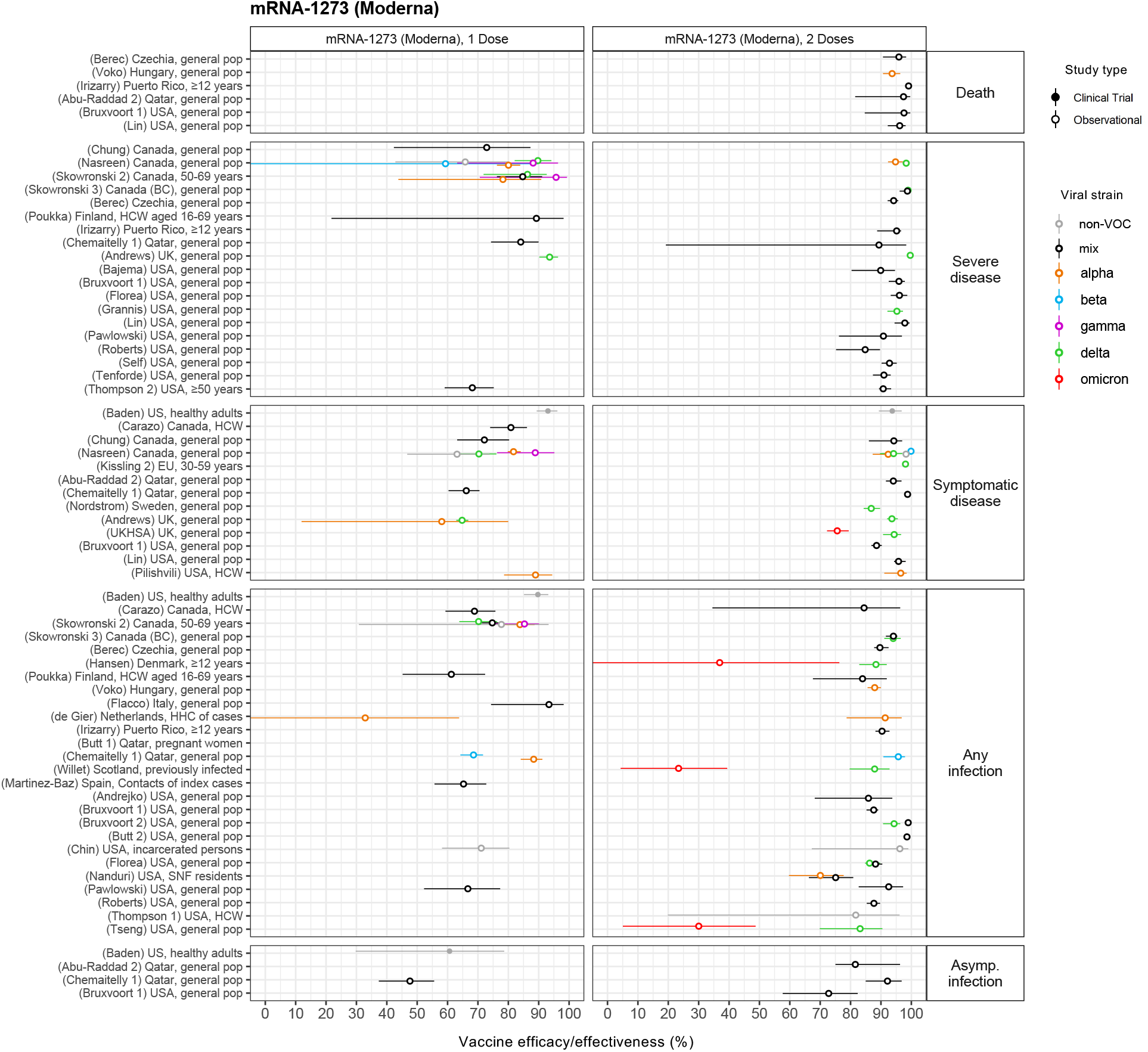
Vaccine efficacy and effectiveness estimates for mRNA-1273, a two-dose mRNA vaccine developed by Moderna. Estimates are colored by the viral variant against which the vaccine efficacy or effectiveness value was measured. Solid markers are estimates from randomized clinical trials (efficacy values), and open markers are estimates from observational studies (effectiveness values). The source of each estimate is given by the labels on the left side (“(reference number) Country, population”). Within each disease severity level, estimates are ordered alphabetically by country, and then by population.

**Figure 5.**
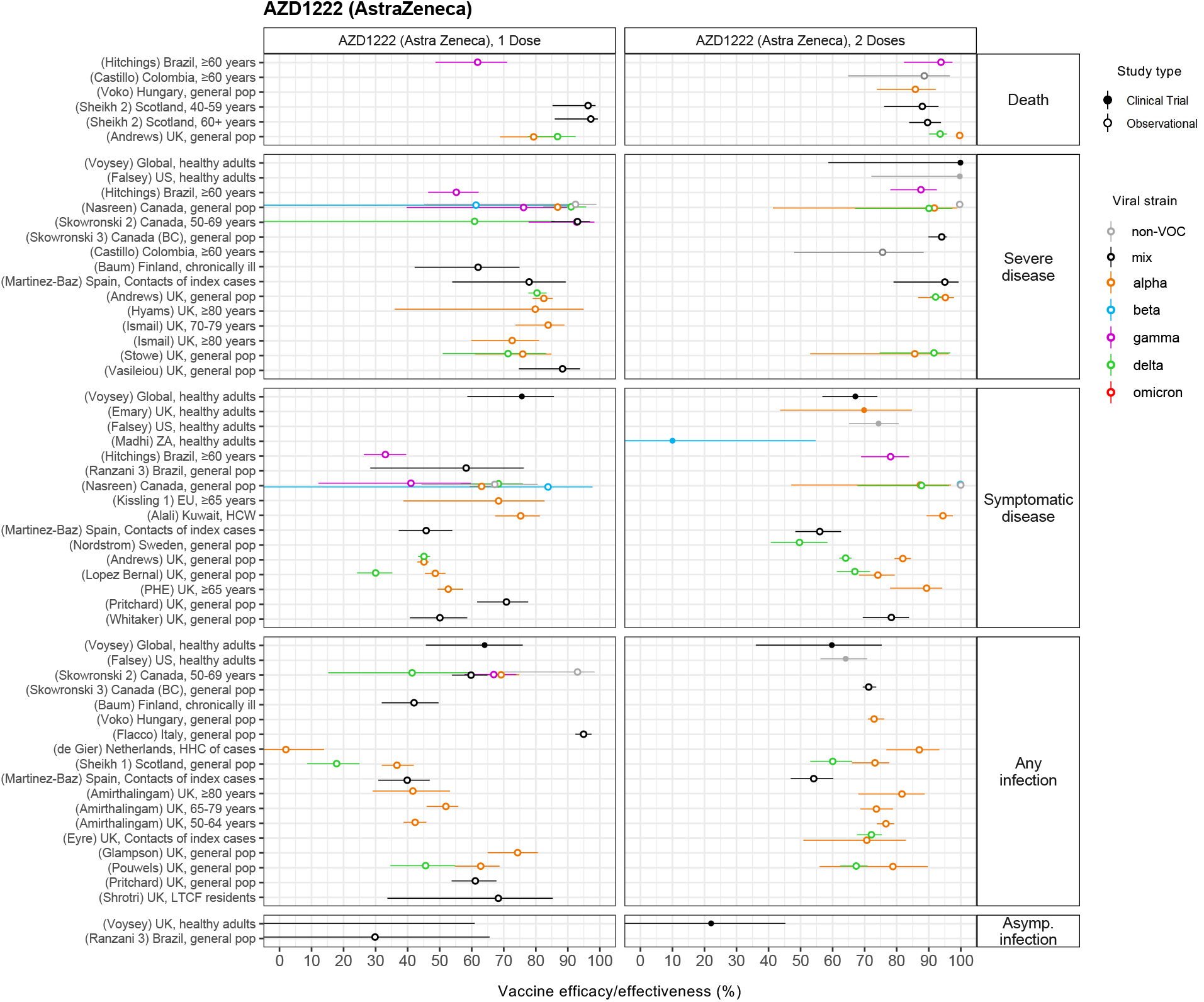
Vaccine efficacy and effectiveness estimates for AZD1222, a two-dose viral vector vaccine developed by AstraZeneca. Estimates are colored by the viral variant against which the efficacy or effectiveness value was measured. Solid markers are estimates from randomized clinical trials (efficacy values), and open markers are estimates from observational studies (effectiveness values). The source of each estimate is given by the labels on the left side (“(reference number) Country, population”). Within each disease severity level, estimates are ordered alphabetically by country, and then by population.

**Figure 6.**
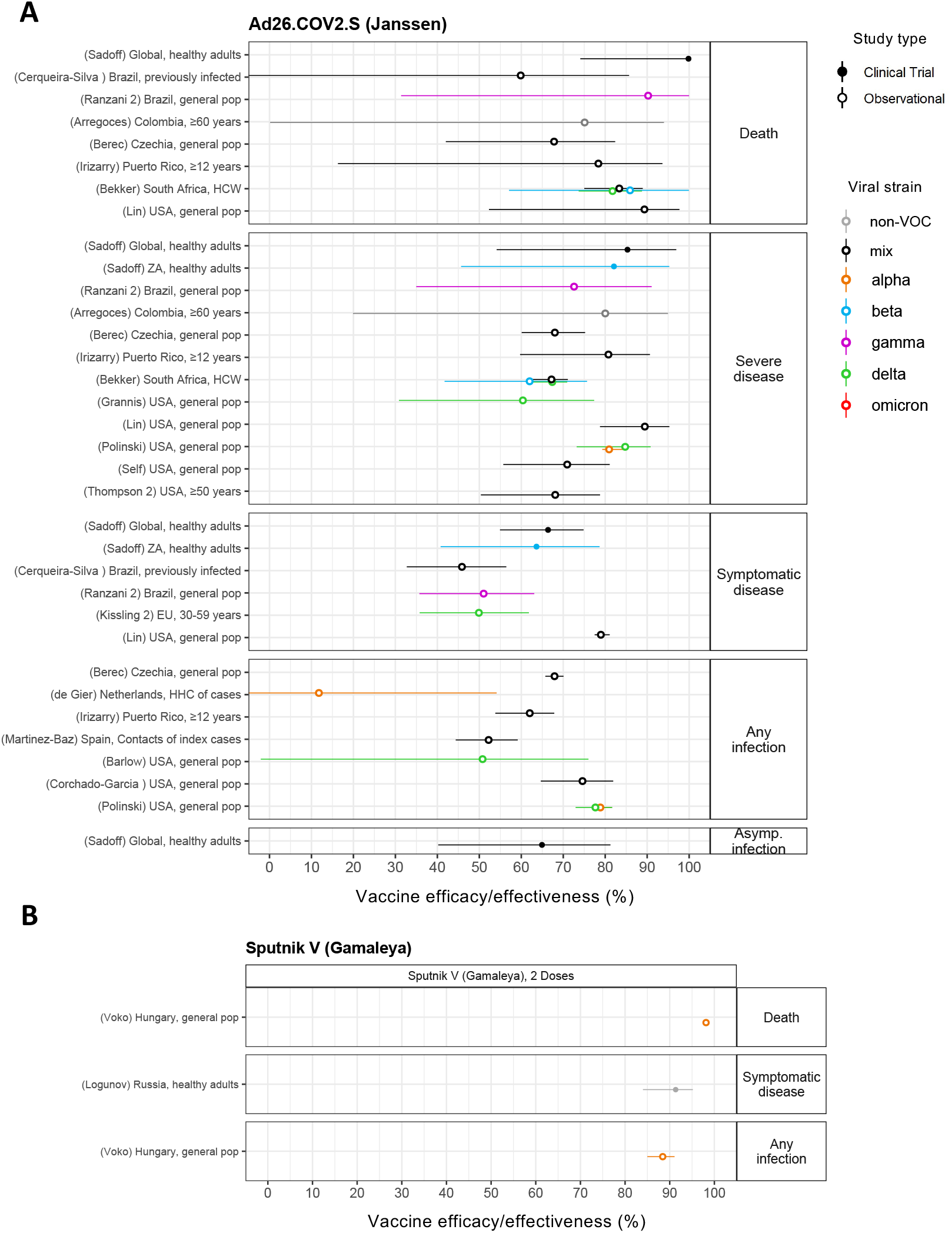
Vaccine efficacy and effectiveness estimates for two viral vector vaccines: A) Ad26.COV2.S, a single-dose vaccine developed by Janssen/Johnson & Johnson, and B) Sputnik V, a two-dose vaccine developed by Gamaleya Institute. Estimates are colored by the viral variant against which the efficacy or effectiveness value was measured. Solid markers are estimates from randomized clinical trials (efficacy values), and open markers are estimates from observational studies (effectiveness values). The source of each estimate is given by the labels on the left side (“(reference number) Country, population”). Within each disease severity level, estimates are ordered alphabetically by country, and then by population.

**Figure 7.**
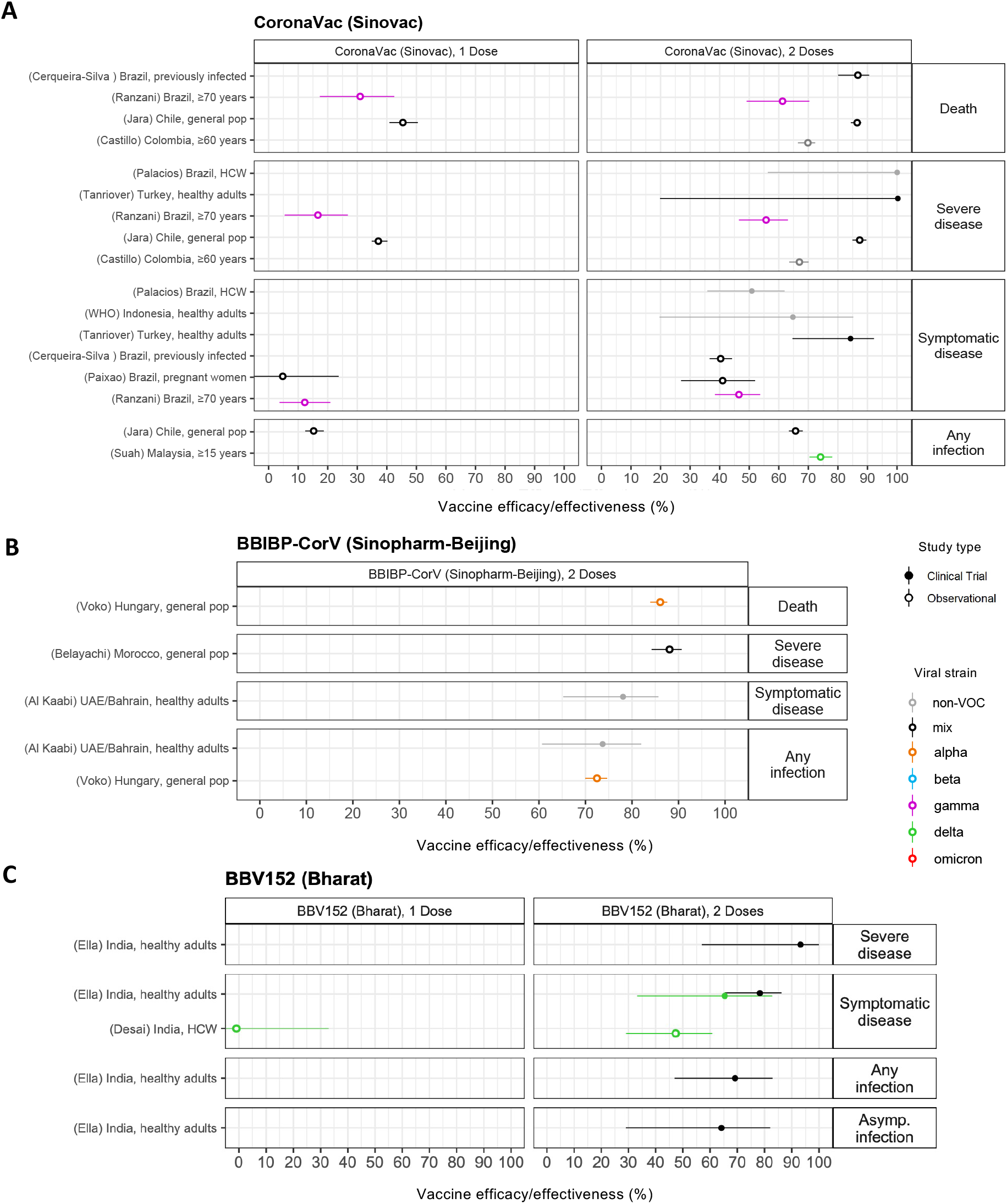
Vaccine efficacy and effectiveness estimates for three two-dose inactivated virus vaccines: A) CoronaVac, developed by Sinovac B) BBIBP-CorV, developed by Sinopharm Beijing, and C) BBV152, developed by Bharat Biotech. Estimates are colored by the viral variant against which the efficacy or effectiveness value was measured. Solid markers are estimates from randomized clinical trials (efficacy values), and open markers are estimates from observational studies (effectiveness values). The source of each estimate is given by the labels on the left side (“(reference number) Country, population”). Within each disease severity level, estimates are ordered alphabetically by country, and then by population.

### Data available from observational studies of vaccine effectiveness

We identified 107 real-world vaccine effectiveness studies meeting our inclusion criteria^4–14,19,26–29,38,84,93–179^. These studies covered eight vaccines: seven with WHO EUL authorization (all but Novavax’s NVX-CoV2373) and one with EUL submission in progress (Gamaleya’s Sputnik V)(**Table 2, <u>Figure S1, Table S1</u>**). These observational studies provided metrics not reported in clinical trials for some vaccines, including effectiveness against severe outcomes, asymptomatic infection, any infection, specific circulating variants, and effectiveness of a single dose of two-dose vaccines (**Figures 3-7**). In general, effectiveness estimates overlapped with efficacy estimates and were high for full vaccine courses. Vaccines were more effective at preventing severe infection or death compared to symptomatic COVID-19. The degree to which vaccines prevented any infection, and the degree to which partial courses prevented infection or disease, varied widely by product. Data were sparse for effectiveness against death or asymptomatic infection and the Gamma variant.

### Efficacy/effectiveness of mRNA vaccines

The most data was available for BNT162b2, the two-dose mRNA vaccine developed by Pfizer/BioNTech, mainly due to its early 2021 use in Israel, the UK, and the US (**Figure 3**). Efficacy/effectiveness estimates after two doses in the general population, pre-Omicron, ranged from 90-99% for death, 80-100% for severe infection, 70-100% for symptomatic disease, 65-98% for any infection, and 65-90% for asymptomatic infection. These values were lower after only a single dose: 70-90% for death, 55-95% for severe infection, 35-93% for symptomatic disease, and 30-80% for any infection. Some studies found lower effectiveness in special populations, including residents of long-term care facilities^84,149,150^, priority groups^122^, the elderly^147^, and those previously infected^113,179^. Heterogeneities between studies made direct comparisons of efficacy/effectiveness for variants difficult. A few studies with head-to-head comparisons suggested slightly reduced effectiveness for BTN162b2 against Beta and Delta compared to Alpha or non-VOC, for symptomatic cases or any infection^13,28,96,123,145,162^. Evidence for reduced effectiveness against Omicron was more pronounced, with protection dropping to ∼ 70% for severe disease, ∼ 65% for symptomatic disease, and ∼55% for any infection^26,27,136^.

The other authorized two-dose mRNA vaccine, Moderna’s mRNA-1273, was also relatively well-studied, largely using data from the USA, Canada, and Qatar (**Figure 4**). After dose 2, efficacy/effectiveness estimates in the general population (pre-Omicron) fell between 94-99% for death, 85-100% for severe disease, 87-100% for symptomatic disease, 83-100% for any infection, and 73-93% for asymptomatic infection. With only a single dose, values were 60-95% for severe disease, 60-95% for symptomatic disease, 65-95% for any infection, and 45-60% for asymptomatic infection. Studies in the UK, USA, and Denmark^27,29,136^ estimated effectiveness against the Omicron variant after two doses was reduced to ∼75% for symptomatic infection and to 30-40% for any infection. Some evidence for reduction in effectiveness against infection or severe disease with Beta after only 1 dose was observed in studies in Canada and Qatar^12,114^. Significant reductions were not consistently reported for effectiveness against other variants of concern.

### Efficacy/effectiveness of viral-vector vaccines

AstraZeneca’s two-dose viral vector vaccine (AZD1222) was also frequently studied, especially after only one dose, likely because the recommended interval between doses is longer than other vaccines (12 weeks) and some countries including the UK and Canada adopted the strategy of prioritizing first doses over second doses in early 2021 (**Figure 5**). Efficacy/effectiveness against non-VOC strains, the Alpha variant (predominant during most studies), or mixes of strains after two doses was 85-100% for death or severe disease, 65-100% for symptomatic disease, 60-80% for any infection, and 22% for asymptomatic infection in the general population. After a single dose, efficacy/effectiveness was 80-96% for death, 75-92% for severe disease, 45-75% for symptomatic disease, and 35-95% for any infection. The Delta variant appeared to reduce effectiveness against symptomatic disease or infection compared to the earlier Alpha variant by 10-15% after 2 doses and ∼20-30% after a single dose in some^13,19,96,145,171^ but not all^12,123^ studies. Effectiveness against the Gamma variant was within the range of earlier strains. Clinical trial evidence from the South Africa site^82^ suggested loss of efficacy against symptomatic infection with the Beta variant, but with much uncertainty (efficacy 10% CI [<0, 55]). No studies meeting our inclusion criteria reported on the protection that AZD1222 alone provided against the Omicron variant.

Janssen’s Ad26.COV2.S single-dose viral vector vaccine was studied in a variety of settings around the world (**Figure 6**), with efficacy/effectiveness estimates of 70-100% against death, 60-90% for severe disease, 50-80% for symptomatic disease, 50-80% for any infection, and 65% for asymptomatic infection. Protection against infection or severe outcomes appeared to be preserved with SARS-CoV-2 variants of concern, although confidence intervals were extremely wide and no data for Omicron were available meeting inclusion criteria.

Data were sparse for Gamaleya Institutes’s two-dose viral vector vaccine Sputnik V (**Figure 6**), despite its use in dozens of countries (**Table 1**). A single clinical trial estimated efficacy against symptomatic disease with non-VOC virus of 91%^72^, while an observational study in Hungary in the context of the Alpha variant reported effectiveness of 98% against death and 88% against any infection^178^. No studies measuring protection against the Beta, Delta, or Omicron variant met our inclusion criteria.

### Efficacy/effectiveness of inactivated virus vaccines

Estimates of vaccine efficacy and effectiveness were available for three different two-dose inactivated virus vaccines: CoronaVac (Sinovac), BBIBP-CorV (Sinopharm-Beijing), and BBV152 (Bharat) (**Figure 7**). Effectiveness studies of Sinovac’s inactivated virus vaccine CoronaVac were available from only six studies, despite being one of the most widely used vaccines worldwide. These complemented results from three separate clinical trials, including one restricted to health care workers. After two doses, in the general population, efficacy/effectiveness was 86% against death, 88-100% against severe disease, 65-85% against symptomatic disease, and 65-75% against any infection. With only one dose, these values were significantly reduced, to 46% for death, 37% for severe disease, and 16% for any infection. Lower values were reported in studies in older adults, including one study conducted in adults over 70 years old in Brazil during a time when Gamma was predominant^163^. The only other variant-specific study suggested effectiveness against any infection was preserved against the Delta variant^180^. Sinopharm-Beijing’s BBIBP-CorV could only be assessed after both doses and using two observational studies ^104,178^ and a single clinical trial^89^. Efficacy/effectiveness 86% for death, 88% for severe disease, 78% for symptomatic disease, and 74% for any infection. The only information on variant-specific protection came from the study from Hungary, conducted during a time when the Alpha variant dominated, and suggested that protection against any infection was similar to non-VOC strains. Bharat’s BBV152 was evaluated in only a single effectiveness study in health care workers^121^ in addition to the initial Phase 3 trial^65^. Efficacy was 93% for severe disease, 78% for symptomatic disease, 69% for any infection and 64% for asymptomatic infection. The RCT suggested that protection against symptomatic disease with the Delta variant was reduced by 10-15 percentage points, while the observational study suggested <50% effectiveness against Delta after 2 doses and none after only 1 dose.

### Efficacy/effectiveness of other vaccines

Although at the time of writing there were at least 10 other COVID-19 vaccines that had received emergency authorization for widespread use in at least one country, we did not identify effectiveness studies meeting our inclusion criteria.

## Discussion

The development of COVID-19 vaccines has been an astounding feat of science. Within a year of detecting the first outbreak and isolating the SARS-CoV-2 virus, multiple vaccines were deployed around the world. In this review, we systematically collected efficacy and effectiveness values by vaccine platform, disease outcome, number of doses, and SARS-CoV-2 variant. These findings demonstrate robust evidence for the high efficacy of COVID-19 vaccines in clinical trials and high effectiveness in real-world settings. We found that across all vaccine platforms, protection against severe infection or death in the general population was at least 60% and most often close to 100%. Efficacy/effectiveness against symptomatic disease was heterogeneous between vaccine products and studies but was almost always greater than 50% and often greater than 90%. The vast majority of studies showed that vaccines provided protection against infection - not just disease - demonstrating the potential for indirect protective effects (“herd immunity”) via reduced transmission. The degree of protection offered by only a single dose of two-dose vaccine courses varied by product. Most vaccines retained high levels of protection for most SARS-CoV-2 variants, especially against severe outcomes. Some studies provided evidence of slight reductions in efficacy/effectiveness for infection or mild disease with the Beta and Delta strains for some vaccines. Although few studies of the Omicron variant were available at publication time, the available evidence supports larger reductions in protection against both mild and severe infection.

There are several important components of COVID-19 vaccine efficacy/effectiveness not addressed here. Vaccine-induced protection can wane over months or years^59,60,78,79,181^. While all estimates reviewed here were restricted to within 6 months of vaccination, waning is a critical issue to monitor for COVID-19 vaccines. A recent meta-analysis estimated efficacy/effectiveness drops ∼10% for severe disease and ∼20% for infection over five months^85^. These drops appear to be more extreme for the Omicron variant; but evidence suggests “booster” doses - administered in many countries during Fall/Winter 2021 - enhance immunogenicity and effectivenes^20,26,27,29,136,182,183^. However, the WHO stresses prioritization of primary doses globally^184^.

Unvaccinated individuals previously infected with SARS-CoV-2 have some protection against re-infection, estimated in recent study to be ∼90%^185,186^. Consequently, including previously-infected persons in the unvaccinated group of studies could bias estimates of effectiveness downward. Alternatively, if prior immunity synergizes with vaccine-induced immunity to enhance protection, this bias could be in the opposite direction. Of 107 observational studies included in this review, 33 are known to have included persons with previous SARS-CoV-2 infection. For studies that stratified efficacy/effectiveness estimates by prior infection status, results were mixed, finding higher, lower, or unchanged values in previously-infected individuals^107,187^.

For multi-dose vaccine regimens, the time interval between doses may affect protection. Efficacy of AZD1222 (AstraZeneca) increased from 55% with <6 weeks between doses to 81% with >12 weeks^3^. In real-world settings, dose interval sometimes varied due to policies of delaying second doses in favor of universal partial vaccination^188,189^, allowing for evaluation of its impact on neutralizing antibody levels and effectiveness^6,19,190^.

While most vaccine manufacturers and international advisory committees (including WHO^191^) initially recommended all doses be with the same product, many countries allowed for “mixing and matching” of doses^192–194^. Effectiveness studies have shown that boosting AZD1222 with either mRNA vaccine increases neutralizing antibody levels compared to two doses of AZD1222 alone^195–199^, and is also more effective against preventing infection^153,171,200^. Further evaluation is needed to understand which COVID-19 vaccine combinations are safe and effective.

For an individual, the goal of vaccination against COVID-19 is to prevent disease, but from a population perspective, there is an additional goal to reduce transmission, which can eliminate the need for complete vaccine coverage. Some studies have observed a reduction in transmission in vaccinated individuals infected with SARS-CoV-2^201^ by testing close contacts of cases^10,123,143,149,162,202–204^. Others have reported reductions in viral load in the respiratory tract - expected to be a determinant of infectiousness - in vaccinated (versus unvaccinated) cases^19,166,176,205^.

Currently, approval of new COVID-19 vaccines typically requires Phase III trials directly measuring vaccine efficacy. However, as vaccine availability increases in the face of an ongoing epidemic, further placebo-controlled trials may be considered impractical or unethical. Instead, new vaccines or immunization schedules will likely be evaluated using “immunobridging”: indirectly inferring vaccine efficacy by measuring immunological biomarkers (like neutralizing antibody levels) identified as correlates of protection^53,55,206–208^. Such study designs - not reviewed here - have already been used for early assessment of waning rates^207,209^, identifying potential loses in protection against new variants^24,25,210^, promoting the need for booster doses^24,25,211,212^, justifying heterologous vaccine courses^212,213^, expanding eligible age groups to children^214,215^, and to grant initial emergency approval to some vaccines (e.g. Medigen’s MVC-COV1901 in Taiwan^216^). As pre-approval efficacy trials become rarer, post-authorization vaccine effectiveness studies will become even more important.

While our review focused on studies conducted in adults, some countries have now expanded vaccination campaigns to include children as young as 2 years old^217,218^. Limited studies suggest that comparable safety, immunogenicity, efficacy, and effectiveness to adults can be achieved with adjusted dosing^219–222^. While children are at lower risk for severe disease, they are susceptible to infection, contribute to transmission, and - because they are more likely to have mild symptoms - may be less likely to be tested or reported as cases, and to isolate during infection. Thus, the ability of COVID-19 vaccines to prevent asymptomatic infection and reduce transmission is especially relevant to the population-level impact of pediatric vaccination.

In conclusion, data from a wide variety of study types and settings demonstrate that COVID-19 vaccines provide high levels of protection against severe disease, and additionally protect against infection and mild disease, even those caused by most SARS-CoV-2 variants of concern. Preliminary evidence supports the idea that the spread of the Omicron variant beginning late 2021 is at least partially driven by reductions in vaccine effectiveness.

## Data Availability

All VE metrics reported in the review are provided in the data supplement (Supplementary Table 1)

## AUTHOR CONTRIBUTIONS

ALH proposed the Review with input from BW, CBJ, JGR, MDK, MMH, and SAT. AAN, ALH, BW, CBJ, and JGR and conducted the search for clinical trials of COVID-19 vaccines and extracted efficacy values. AB, PSZ, KW and MMH conducted the search for observational studies of COVID-19 vaccines and extracted effectiveness values. MDK, DRF, and MKP supervised the collection of vaccine effectiveness studies including designing the search strategy, choosing the inclusion criteria, and evaluating studies against these criteria. ALH and MMH synthesized and interpreted the data on efficacy and effectiveness. BW assembled data on clinical trial timing, location, and SARS-CoV-2 variant prevalence. ALH, BW, and MMH created the tables and figures. ALH, CBJ, BW, JGR and MHH drafted the manuscript. All authors revised the manuscript and approved it for submission.

## ACKNOWLEDGMENTS

The authors thank M Kate Grabowski and the Johns Hopkins University Novel Coronavirus Research Compendium for bringing the study authors together to work on this paper.

## PATIENT CONSENT STATEMENT

This study did not include factors necessitating patient consent

