## Supplementary Material for "A systematic review of COVID-19 vaccine efficacy and effectiveness against SARS-CoV-2 infection and disease"

### Supplementary Methods

#### Notes on figures

Figure 1: We included countries that had both a) available data on SARS-CoV-2 variants of concern over time reported to GISAID, and b) locations of clinical trials for COVID-19 vaccines in the WHO EUL pipeline that had at least 10,000 participants from the general healthy adult population. For example, we did not show Sinovac's trial of CoronaVac since it only included health care workers nor Pfizer's trial locations in Brazil, South Africa, or Argentina due to small sizes. We did not plot the BioCubaFarma or Sinopharm trials since no viral sequence data was available for Cuba or the United Arab Emirates. Since variant prevalences were only reported for aggregated two week periods on GISAID, we reported the average daily incidence of reported cases for the same two week periods by aggregating daily data. Incidence was normalized by the country's population. The start of the clinical trial observation period was defined as the earliest time point two weeks after first dose of vaccination until the end date of cases included in the analysis. These were estimated from the protocols/supplementary material of the publications for each trial.

Figure 2: We chose a single efficacy metric to report for each vaccine. If there were multiple clinical trial sites, we plotted the pooled estimate if it was available or the one from the largest trial done in the general healthy adult population. For AstraZeneca (AZD1222), we reported the value pooled over all inter-dose timings. For Bharat (BBV152) we reported the overall efficacy against all SARS-CoV-2 strains. For Janssen (Ad26.COV2.S) and Pfizer (BNT162b2) we reported the pooled efficacy over all study sights ("global"). For Novavax (NVX-CoV2373) we reported the value for the US/Mexico trial since it was the largest. For Sinovac (CoronaVac) we reported the value for the trial in Turkey, since it was approximately equal in size to the trial in Brazil but involved the general adult population instead of only health care workers.

Figures 3-7: Within each clinical outcome group, points were ordered first whether the study was randomized vs observational, secondly alphabetically by country of study, and thirdly alphabetically by first author surname. An estimate was not included in the plots if a more recent estimate from an updated analysis/publication for the same population was available. In addition, if a study reported estimates for the same population using two different methodologies, only the estimate from the primary methodology was selected. For studies providing estimates stratified by prior SARS-CoV-2 infection status, the estimate excluding previously infected persons was selected.

#### Vaccine effectiveness literature search terms

Overview: Search criteria were run every week from February 1, 2021 to February 3, 2022, and all new reports published since the past search were reviewed. The International Vaccine Access Center at Johns Hopkins University conducted these searches and reviews on behalf of the WHO from June 17, 2021 to February 3, 2022. Their reviews, and the results included here, additionally include the results of searches that were conducted from February 1 to June 17,

2021 by the CDC, with slightly different search methods. Both search strategies are described below. In addition to these searches, all pre-prints on the medRxiv including the terms “COVID” or “SARS-CoV-2” in the title were reviewed manually.

Current search strategy (June 17, 2021 - February 3, 2022)

PubMed:

("COVID-19"[tw] OR "COVID 19"[tw] OR "COVID19"[tw] OR "COVID2019"[tw] OR "COVID 2019"[tw] OR "COVID-2019"[tw] OR "novel coronavirus"[tw] OR "new coronavirus"[tw] OR "novel corona virus"[tw] OR "new corona virus"[tw] OR "SARS-CoV-2"[tw] OR "SARSCoV2"[tw] OR "SARS-CoV2"[tw] OR "2019nCoV"[tw] OR "2019-nCoV"[tw] OR "2019 coronavirus"[tw] OR "2019 corona virus"[tw] OR "coronavirus disease 2019"[tw] OR "severe acute respiratory syndrome coronavirus 2"[nm] OR "severe acute respiratory syndrome coronavirus 2"[tw] OR "sars-coronavirus-2"[tw] OR "coronavirus disease 2019"[tw] OR "corona virus disease 2019"[tw])
AND
("COVID-19 Vaccines"[Mesh] OR “COVID-19 vaccine”[tiab] OR "mRNA-1273 vaccine" [Supplementary Concept] OR “mRNA-1273 vaccine”[tiab] OR “mRNA vaccine”[tiab] OR “mRNA COVID-19 vaccines”[tiab] OR "ChAdOx1 COVID-19 vaccine" [Supplementary Concept] OR "Ad5-nCoV vaccine" [Supplementary Concept] OR “Ad5-nCoV”[tiab] OR "Covid-19 aAPC vaccine" [Supplementary Concept] OR "Ad26.COV2.S vaccine" [Supplementary Concept] OR "Ad26.COV2.S vaccine"[tiab] OR “adenoviral vector vaccine”[tiab] OR "BNT162 vaccine" [Supplementary Concept] OR “BNT162b2”[tiab] OR “BNT162”[tiab] OR “CoronaVac” [tiab] OR “vaccin*”[tiab])
AND
("Clinical Trial, Phase IV" [Publication Type] OR "Controlled Clinical Trial" [Publication Type] OR "Randomized Controlled Trial" [Publication Type] OR "Case-Control Studies"[Mesh] OR "Retrospective Studies"[Mesh] OR “Retrospective”[tiab] OR "Cohort Studies"[Mesh] OR "Prospective Studies"[Mesh] OR “Prospective”[tiab] OR "Longitudinal Studies"[Mesh] OR "Follow-Up Studies"[Mesh] OR “Follow-up studies”[tiab] OR “cohort”[tiab] OR "test negative"[tiab] OR “Observational cohort”[tiab] OR “Test-negative design”[tiab] OR"RCT"[tiab] OR "randomized"[tiab] OR "randomised"[tiab] OR "randomly allocated"[tiab] OR “case-control”[tiab] OR “real-world effectiveness”[tiab] OR “effectiveness”[tiab] OR “association”[tiab] OR “impact”[tiab] OR “vaccine impact”[tiab])
NOT ("Clinical Trial, Phase I" [Publication Type] OR "Clinical Trial, Phase II" [Publication Type])
NOT (“animals”[mesh] NOT (“animals”[mesh] AND “humans”[mesh]))

Embase:

('COVID-19' OR 'COVID 19' OR 'COVID19' OR 'COVID2019' OR 'COVID 2019' OR 'COVID-2019' OR 'novel coronavirus' OR 'new coronavirus' OR 'novel corona virus' OR 'new corona virus' OR 'SARS-CoV-2' OR 'SARSCoV2' OR 'SARS-CoV2' OR '2019nCoV'

OR '2019-nCoV' OR '2019 coronavirus' OR '2019 corona virus' OR 'coronavirus disease 2019' OR 'severe acute respiratory syndrome coronavirus 2'/exp OR 'severe acute respiratory syndrome coronavirus 2' OR 'sars-coronavirus-2' OR 'coronavirus disease 2019'/exp OR 'coronavirus disease 2019' OR 'corona virus disease 2019')
AND
('SARS-CoV-2 vaccine'/exp OR 'COVID-19 vaccine':ti,ab OR 'mRNA-1273 vaccine'/exp OR 'mRNA-1273 vaccine':ti,ab OR 'mRNA vaccine':ti,ab OR 'mRNA COVID-19 vaccines':ti,ab OR 'ChAdOx1 ncov 19'/exp OR 'Ad5 nCoV vaccine'/exp OR 'Ad5-nCoV':ti,ab OR 'Covid-19 aAPC vaccine':ti,ab OR 'Ad26.COV2.S vaccine'/exp OR 'Ad26.COV2.S vaccine':ti,ab OR 'adenoviral vector vaccine':ti,ab OR 'BNT 162 vaccine'/exp OR 'BNT162b2':ti,ab OR 'BNT162':ti,ab OR 'CoronaVac'/exp OR 'coronavac':ti,ab OR 'vaccin*':ti,ab)
AND
('phase 4 clinical trial'/exp OR 'Controlled Clinical Trial'/exp OR 'Randomized Controlled Trial'/exp OR 'Case Control Study'/exp OR 'Retrospective Study'/exp OR 'Retrospective':ti,ab OR 'Cohort analysis'/exp OR 'Prospective Study'/exp OR 'Prospective':ti,ab OR 'Longitudinal Study'/exp OR 'Follow Up'/exp OR 'Follow-up study':ti,ab OR 'cohort':ti,ab OR 'test negative':ti,ab OR 'Observational cohort':ti,ab OR 'postmarketing surveillance'/exp OR 'postmarketing surveillance':ti,ab OR 'Test-negative design':ti,ab OR 'RCT':ti,ab OR 'randomized':ti,ab OR 'randomised':ti,ab OR 'randomly allocated':ti,ab OR 'case-control':ti,ab OR 'real-world effectiveness':ti,ab OR 'effectiveness':ti,ab OR 'association':ti,ab) NOT ('phase 1 clinical trial'/exp OR 'phase 2 clinical trial'/exp)
NOT ('animal'/exp NOT ('animal'/exp AND 'human'/exp))
NOT 'conference abstract'/it

<u>WHO COVID Global Literature Database:</u>

("COVID-19 Vaccines" OR "COVID-19 vaccine" OR "mRNA-1273 vaccine" OR "mRNA vaccine" OR "mRNA COVID-19 vaccines" OR "ChAdOx1 COVID-19 vaccine" OR "Ad5-nCoV" OR "Covid-19 aAPC vaccine" OR "Ad26.COV2.S vaccine" OR "adenoviral vector vaccine" OR "BNT162b2" OR "BNT162" OR "CoronaVac" OR vaccin*)
AND
("Phase IV" OR "Controlled Clinical Trial" OR "Randomized Controlled Trial" OR "Case-Control Studies" OR "Retrospective" OR "Cohort Studies" OR "Prospective" OR "Longitudinal Studies" OR "Follow-Up Studies" OR "Follow-up study" OR "cohort" OR "test negative" OR "Observational cohort" OR "Test-negative design" OR "RCT" OR "randomized" OR "randomised" OR "randomly allocated" OR "case-control" OR "real-world effectiveness" OR "effectiveness" OR "association") AND NOT ("Phase I" OR "Phase II")

<u>SCOPUS:</u>

TITLE-ABS-KEY(“novel coronavir*” OR “novel corona virus*” OR “2019 coronavirus” OR betacoronavir* OR covid19 OR "covid 19" OR ncov OR "CoV 2" OR cov2 OR sarscov2 OR sars-cov OR sarscov OR 2019ncov OR 2019-nCoV OR "novel CoV" OR “coronavirus infections”) AND TITLE-ABS-KEY(Vaccin* AND (effectiveness OR efficacy OR protection*) AND (postmarketing OR approved OR (post* W/5 approval) OR "real world" OR "phase IV" OR "phase 4" OR observational OR longitudinal OR spread OR transmission OR (rate* W/5 infection*) OR (reduc* W/5 infection*) OR "general population"))

Web of Science:

(TI=(covid-19 vaccine effectiveness )) OR AB=(covid-19 vaccine effectiveness )

medRxiv, bioRxiv, SSRN, Europe PMC, Research Square, Knowledge Hub:

“COVID-19 vaccine effectiveness” OR “COVID-19 vaccine efficacy”

In addition to the above databases, MMWR and Eurosurveillance are hand-searched weekly for new studies meeting eligibility criteria.

Prior search strategy (February 1, 2021 - June 17 2021)

Medline (via OVID):

(Vaccin* AND (effectiveness OR efficacy OR protection*) AND (postmarketing OR approved OR (post* ADJ5 approval) OR real world OR phase IV OR phase 4 OR observational OR longitudinal OR spread OR transmission OR (rate* ADJ5 infection*) OR (reduc* ADJ5 infection*) OR general population))
Limit to COVID-19

Coronavirus Research Database (via ProQuest) AND ProQuest Central

TI,AB(“novel coronavir*” OR “novel corona virus*” OR “2019 coronavirus” OR betacoronavir* OR covid19 OR “covid 19” OR nCoV OR “novel CoV” OR “CoV 2” OR CoV2 OR sarscov2 OR sars-cov OR sarscov OR 2019nCoV OR 2019-nCoV OR “coronavirus infections”)
AND
TI,AB(Vaccin* AND (effectiveness OR efficacy OR protection*) AND (postmarketing OR approved OR (post* NEAR/5 approval) OR "real world" OR "phase IV" OR "phase 4" OR observational OR longitudinal OR spread OR transmission OR (rate* NEAR/5 infection*) OR (reduc* NEAR/5 infection*) OR "general population"))

Scopus:

TITLE-ABS-KEY(“novel coronavir*” OR “novel corona virus*” OR “2019 coronavirus” OR betacoronavir* OR covid19 OR "covid 19" OR ncov OR "CoV 2" OR cov2 OR sarscov2 OR sars-cov OR sarscov OR 2019ncov OR 2019-nCoV OR "novel CoV" OR “coronavirus infections”) AND TITLE-ABS-KEY(Vaccin* AND (effectiveness OR efficacy OR protection*) AND (postmarketing OR approved OR (post* W/5 approval) OR "real world" OR "phase IV" OR "phase 4" OR observational OR longitudinal OR spread OR transmission OR (rate* W/5 infection*) OR (reduc* W/5 infection*) OR "general population"))

WHO COVID Global Literature Database:

(tw:(vaccine OR vaccines OR vaccination OR vaccinations)) AND (tw:(effectiveness OR efficacy OR protection OR protections)) AND (tw:(postmarketing OR approved OR approval OR "real world" OR "phase IV" OR observational OR longitudinal OR spread OR transmission OR "phase 4" OR "general population"))

CINAHL (via EbscoHost):

(“novel coronavir*” OR “novel corona virus*” OR “2019 coronavirus” OR betacoronavir* OR covid19 OR “covid 19” OR nCoV OR “novel CoV” OR “CoV 2” OR CoV2 OR sarscov2 OR sars-cov OR sarscov OR 2019nCoV OR 2019-nCoV OR “coronavirus infections”)
AND
(Vaccin* AND (effectiveness OR efficacy OR protection*) AND (postmarketing OR approved OR (post* N5 approval) OR "real world" OR "phase IV" OR "phase 4" OR observational OR longitudinal OR spread OR transmission OR (rate* N5 infection*) OR (reduc* N5 infection*) OR "general population"))

### Supplementary Tables

**Table S1: List of all study characteristics and individual vaccine efficacy and effectiveness metrics included in the review.** Attached separately as an Excel file.

### Supplementary Figures

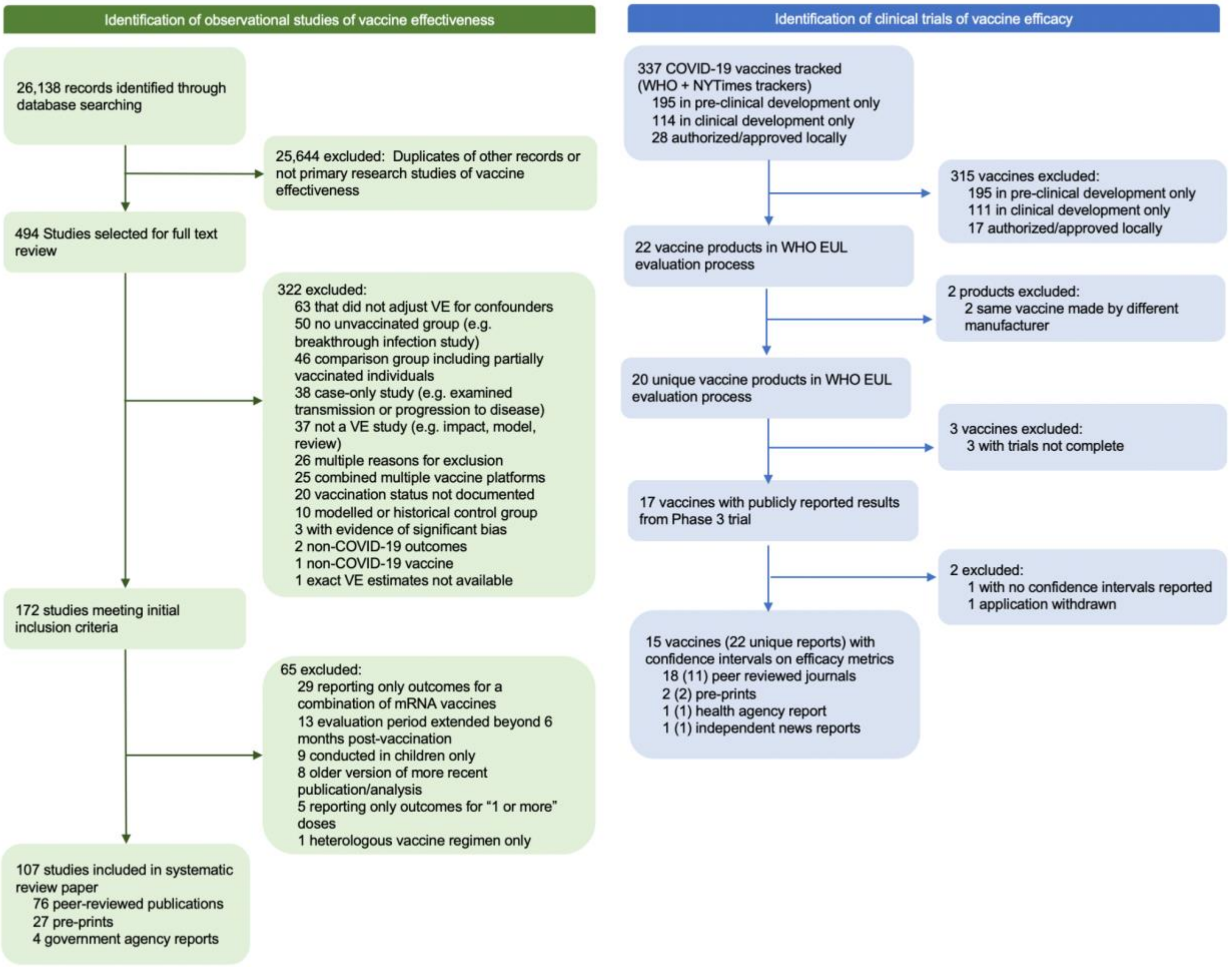


**Figure S1. PRISMA flow diagram for systematic review of COVID-19 vaccine efficacy and effectiveness**
